# Phenotypic and genetic factors associated with differential consent to record linkage for prescription history in the Australian Genetics of Depression Study

**DOI:** 10.1101/2021.10.26.21265507

**Authors:** Lina Gomez, Santiago Díaz-Torres, Lucía Colodro-Conde, Luis M. Garcia-Marin, Chloe Yap, Enda M. Byrne, Loic Yengo, Penelope A. Lind, Naomi R. Wray, Sarah E. Medland, Ian B. Hickie, Michelle K. Lupton, Miguel E. Rentería, Nicholas G. Martin, Adrian I. Campos

## Abstract

Samples can be prone to ascertainment and attrition biases.The Australian Genetics of Depression Study is a large publicly recruited cohort (n=20,689) established to increase the understanding of depression and antidepressant treatment response. As part of the recruitment, participants donated a saliva sample and were given the option to consent to linkage of prescription records for research purposes. This study investigates differences between participants who donated a saliva sample or agreed to linkage of their records compared to those who did not. We observed that older, male participants with a higher education were more likely to donate a saliva sample. Self-reported bipolar disorder, ADHD, panic disorder, PTSD, substance use disorder and social anxiety disorder were associated with lower odds of donating a saliva sample whereas anorexia was associated with higher odds of donation. Male and younger participants showed higher odds of agreeing to record linkage. Participants with higher neuroticism scores and those with a history of bipolar disorder were also more likely to agree to record linkage whereas participants with a diagnosis of anorexia were less likely to agree. Increased likelihood of consent was also associated with increased genetic susceptibility to anorexia and reduced genetic risk for depression, and schizophrenia whereas there was no significant genetic effect for neuroticism. Overall, our results show moderate differences among these subsamples. Most current epidemiological studies do not adjust, nor search, for attrition biases at the genetic level. The possibility to do so is a strength of samples such as the AGDS. Our results suggest that analyses can be made more robust by identifying attrition biases both on the phenotypic and genetic level, and either contextualising them as a potential limitation or performing sensitivity analyses adjusting for them.

## Introduction

The Australian Genetics of Depression Study (AGDS) is a large cohort study including more than 20,000 participants. Recruitment targeted individuals who had been diagnosed or treated for depression [1]. The majority (75%) of the participants in the study are women. The mean age (at the time of recruitment) was 43 years with a standard deviation of 15 years. A high percentage of the participants (95%) have been diagnosed with depression, and similar to what is seen in population studies, 68% of the cohort reported a history of at least one other comorbid mental health diagnosis. The AGDS collected a vast amount of phenotypic data through online questionnaires, as well as a biological specimen for genotyping. A key component of the study was the optional consent to linkage of participants’ prescription history with the study data. These datasets enable a variety of novel analyses such as corroboration of medication self-reports as well as health economics and comorbidity analyses.

Public recruitment practices can be prone to biases as they indirectly target a portion of the population that consists of individuals that are willing to participate in the study and that decision could lead to an over or under representation (relative to the population) of certain traits [2], and even specific disorders [3] that correlate with willingness to participate. A similar phenomenon may occur among participants in their completion of optional modules or participation in subsequent waves of data collection. We refer to this phenomenon as attrition. Previous studies have shown that attrition can influence measures of association [4] by creating a bias in the prevalence and incidence of the variables studied. An example of this is *collider bias*, an artificial association between two variables as the product of either adjusting for a covariate that is actually an outcome of the two variables studied [5], or sampling strategies which modify the likelihood of recruiting participants with specific values for the common outcome [6].

Large cohorts and genetic studies are not free of sampling and attrition biases. Evidence of such include the observation of unexpected autosomal heritability of sex in cohorts with active recruitment, which could possibly reflect differences in genetic factors driving participation in males and females [7], and genetic factors associated with participating in optional subsections of surveys of the UK-Biobank [8,9], all of which maylead to incorrect inferences in downstream analyses. Identifying and quantifying selection biases in cohorts and subsamples is necessary to identify potentially spurious findings, or to account for them statistically where possible. The objective of this study was to evaluate selection biases in two aspects of participation in the ADGS: donating a saliva sample for genotyping and agreeing to linkage to medical prescription records (PBS linkage) in the AGDS. We did this by investigating sociodemographic and psychiatryc differences between participants who donated a saliva sample compared to those who did not. We further tested whether there is evidence for heritability to agree to PBS linkage and investigated sociodemographic, clinical and polygenic differences between participants who agreed to PBS linkage and those who did not. Finally, we compare our results across the two participation measures and with studies of participation in other cohorts such as the UK-Biobank.

## Methods

### Sample recruitment

The Australian Genetics of Depression Study recruited 22,424 Australian participants through two avenues: a mail-out to patients who had at least four prescriptions for antidepressants in the previous 5 years (14%) and media campaign to recruit patients who had received a diagnosis of depression from a doctor, psychiatrist or psychologist (86%). Potential participants were directed to the AGDS website (https://www.geneticsofdepression.org.au), and informed consent was gathered prior to data collection through online questionnaires. The AGDS inclusion criteria included (i) reporting that they had been treated for depression by a health professional (ii) agreeing to donate a saliva sample for genotyping (although only 72.5% actually did so). Full details for the AGDS can be found elsewhere [1].

Among the phenotypes collected, participants confirmed whether they had taken any of the ten most commonly prescribed antidepressants in Australia. For each antidepressant taken, we gathered data on antidepressant efficacy and experienced side effects. Data on demographics, clinical history of psychiatric disorders as well as personality traits (neuroticism and extraversion scores) were also collected. Furthermore, willing participants provided optional consent to record linkage of their Pharmaceutical Benefits Scheme (PBS) prescription and Medical Benefits Scheme (MBS) records, which could be used to validate medication self-reported data, assess concurrent medications and infer comorbidities.

In this study, we focus on 1) whether participants agreed to record linkage of their PBS records for research purposes as the outcome of interest, and 2) whether they donate a saliva sample for DNA genotyping. The full list and details of instruments used for AGDS phenotyping are available at https://bit.ly/3y72lyg. All participants provided informed consent prior to participating in the study. The QIMR Berghofer Medical Research Institute Human Research Ethics Committee approved all questionnaires and research procedures for the AGDS under project number P2118.

### Genotyping imputation and quality control

Upon completion of the core questionnaire, participants were mailed a GeneFix GFX-02 2mL saliva DNA extraction kit (Isohelix plc) to use at home and then returned by mail for subsequent genotyping. The AGDS sample was genotyped using the Illumina Global Screening Array (GSA V.2.0). Genotype data were cleaned by removing unknown or ambiguous map position, strand alignment, high missingness (>5%), deviation from Hardy-Weinberg equilibrium, low minor allele frequency (<1%), and GenTrain score <0.6 variants. Imputation was performed through the Michigan imputation server web service using the HRCr1.1 reference panel, as the majority of the cohort were of European ancestry. Genotyped individuals were excluded from polygenic risk score (PRS) analyses based on high genotype missingness, inconsistent and unresolvable sex, or if deemed ancestry outliers from the European population, based on principal components derived from the 1000Genomes reference panel (defined as >6SD from the PC1/PC2 centroid).

### SNP-based heritability

We employed genome-based restricted maximum likelihood (GREML) as implemented in GCTAv.1.91.7 [10] to estimate the proportion of variance in consent to record linkage (on the observed scale) explained by measured genetic differences (*SNP-based heritability*). This approach leverages a genetic-relatedness matrix and restricted maximum likelihood to partition the variance of a phenotype into a genetic and environmental component [11]. The underlying logic is to assess whether genetic covariation explains a significant proportion of the covariation of a trait. For this study, a GRM based on a subset of unrelated (genomic relatedness cutoff < 0.05) individuals of European ancestry was employed to identify evidence for a genetic component to consenting record linkage of PBS records.

### Polygenic risk scores (PRS)

We computed PRS in order to test to what extent the genetic risk for traits that showed a phenotypic association, or have previously been linked to attrition in other studies (see discussion), predicted consent to PBS record linkage. For the PRS predictions, we excluded European ancestry outliers and used only one member from groups of related individuals (genomic relatedness cutoff < 0.05) to avoid confounding from cryptic relatedness, as this violates the independence assumption of a classical logistic regression. We estimated PRS for educational attainment [12], neuroticism [13], major depressive disorder [14], bipolar disorder [15], schizophrenia (SCZ) [16], and anorexia nervosa [17] using GWAS summary statistics without sample overlap with the AGDS cohort. SBayesR was used to estimate the joint GWAS effect sizes adjusting for the correlation between SNPs [18]. Prior to estimating PRS, we excluded low imputation quality (r^2^ < 0.6), MAF < 0.01, non-autosomal and strand-ambiguous variants. Imputed genotype dosage data were used to calculate PRS by multiplying the variant effect size by the dosage of the effect allele. Finally, the total sum was calculated across all variants. This procedure was performed using Plink 1.9 [19]. A relevant follow up question is how much of the *SNP-based heritability* is explained jointly by these genetic factors. To estimate the proportion of *SNP-based heritability* explained by these PRS, a secondary GREML analysis including all nominally significant PRS (SZC, MDD and anorexia) as a fixed effect was performed, and the SNP based heritability of both models (with and without PRS) were compared using the formula 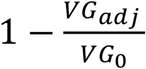 where VG_0_ is the genetic variance component estimated in the null model (i.e., without PRS) and VG_adj_ is the same estimate but of the model including PRS for SCZ, MDD and anorexia as fixed effects. This approach is likely to yield an underestimate as the GRM used to estimate SNP-based heritability is constructed using SNPs also included in the PRS.

### Statistical analyses

Logistic regression was used to examine the association between consent to record linkage (yes/no) and variables of interest including age, sex, educational attainment, psychopathology, and PRS. The non-genetic regressions were adjusted for sex and age at study enrollment. Regressions using genetic variables included age, sex and the first 20 genetic principal components to adjust for potential population stratification. Variance explained was estimated by the regression coefficient of determination squared (R^2^). Results assessing the proportion of *SNP-based heritability* used linear mixed effects models fit via restricted maximum likelihood. Nominally significant results are defined as those with p<0.05, and statistical significance was defined after Bonferroni correction for multiple testing.

## Results

### Demographic factors and samples description

**Table 1** shows demographic information across AGDS participants who did or did not consent to record linkage. We also contrast demographic factors between participants who donated a saliva sample for genotyping and those who did not. Participants agreeing to record linkage consent were, on average, older (OR=1.016; 95%C.I.=[1.01-1.02] per year of age). Female participants were less likely to provide consent for record linkage (OR=0.69; 95%C.I.=[0.63-0.74]). Overall the cohort’s educational attainment is high (e.g. ∼25% of participants reported having a postgraduate degree). We did not observe a significant association between educational attainment and providing consent for record linkage (OR=1.02; 95%C.I.=0.99-1.05). Being male (OR=1.13; 95%C.I.=[1.05-1.23]), older (OR=1.012; 95%C.I.=[1.010-1.015] per year of age) and having a higher educational attainment (OR=1.18; 95%C.I.=[1.14-1.21] per educational category) were all associated with donating a saliva sample for genotyping.

**Table 1.** Relationship between demographics and consent for record linkage and provision of a genetic sample.

|  |  |  |  |  |  |
| --- | --- | --- | --- | --- | --- |
|  | **Whole sample** | **Record linkage** | **No record linkage** | **Biological Sample** | **No biological sample** |
| **Age in years** |  |  |  |  |  |
| **Mean (SD)** | 43.17 (15.42) | 44.3 (15.26) | 39.95 (15.42) | 43.7 (15.3) | 41.9 (15.7) |
| **Sex** |  |  |  |  |  |
| **Female** | 16,790 (75%) | 12,072 (72%) | 4,718 (28%) | 12,166(72.5%) | 4,624 (27.5%) |
| **Male** | 5,474(24,5%) | 4,383 (80%) | 1,091 (20%) | 3,937(71.9%) | 1,537(28.1%) |
| **Education** |  |  |  |  |  |
| **No formal education** | 12 (0.06%) | 7 (0.05%) | 5 (0.01%) | 7 (0.04%) | 5 (0.11%) |
| **Primary school** | 53 (0.3%) | 44 (0.3%) | 9 (0.2%) | 38 (0.24%) | 15 (0.36%) |
| **Junior secondary school** | 1162 (5.75%) | 893 (5.9%) | 269 (5.3%) | 862 (5.4%) | 300 (7.2%) |
| **Senior secondary school** | 1714 (8.5%) | 1232 (8.1%) | 482 (9.6%) | 1252 (7.8%) | 462 (11.1%) |
| **Certificate or diploma** | 4793 (23.7%) | 3692 (24.3%) | 1101 (21.9%) | 3755 (23.4%) | 1038 (24.9%) |
| **Degree** | 7055 (34.9%) | 5206 (34.3%) | 1849 (36.8%) | 5617 (35.1%) | 1438 (34.5%) |
| **Postgraduate degree** | 5399 (26.7%) | 4088 (27.0%) | 1311 (26.1%) | 4485 (28.0%) | 914 (21.9%) |

### Associations with psychiatric and personality traits

The association between self-reported lifetime psychiatric diagnosis and differential consent for record linkage or biological sample are shown in **Table 2**. Participants with self-reported diagnosis of bipolar disorder (OR=1.29 95%C.I[1.16-1.44]) or higher neuroticism score (OR=1.02 95%C.I.=[1.01-1.04]) were more likely to consent to record linkage. In contrast, participants who reported a diagnosis of anorexia (OR=0.68 95%C.I.=[ 0.60 - 0.77]) were less likely to consent to record linkage. There was nominal association between consent to record linkage and participants reporting a lifetime diagnosis of personality disorder (OR= 1.19 95% C.I.=[1.05 - 1.36], substance use disorder (OR= 1.26 95% C.I.=[1.06 - 1.49] or anxiety disorder (OR=1.07 95% C.I.=[1.01 - 1.14]). For the outcome of donating a saliva sample, the following were associated with lower odds of donation, bipolar disorder (OR= 0.83 95% C.I.=[0.75 - 0.92]), ADHD (OR= 0.73 95% C.I.=[0.63 - 0.85]), panic disorder (OR= 0.75 95% C.I.=[0.68 - 0.83]), PTSD (OR= 0.81 95% C.I.=[0.74 - 0.88]), substance use disorder (OR= 0.78 95% C.I.=[0.66 - 0.9]) and social anxiety disorder (OR= 0.93 95% C.I.=[0.88 - 0.99]). Conversely, participants reporting a lifetime diagnosis of anorexia showed higher odds (OR=1.84 95%C.I.=[01.81-2.15]) of providing a saliva sample.

**Table 2.** Association between psychiatric traits and PBS consent.

| Disorders | Consent to record linkage OR (95% C.I.) | Consent to record linkage p value | Biological sample OR (95% C.I.) | Biological sample p value | N cases |
| --- | --- | --- | --- | --- | --- |
| Bipolar Disorder | 1.29 (1.16 - 1.44) | 4.63E-06* | 0.83 (0.75-0.92) | 2.5E-4* | 2057 |
| Anxiety Disorder | 1.08 (1.01 - 1.15) | 1.76E-02 | 0.93 (0.88-0.99) | 0.02 | 12131 |
| Schizophrenia | 1.17 (0.83 - 1.64) | 0.36 | 0.79 (0.58-1.06) | 0.12 | 195 |
| ADHD | 1.04 (0.90 - 1.21) | 0.58 | 0.73 (0.63-0.85) | 1.9E-05* | 900 |
| Agoraphobia | 0.95 (0.77 - 1.16) | 0.62 | 0.8 (0.66-0.96) | 0.02 | 488 |
| Premenstrual dysphoric disorder | 0.99 (0.81 - 1.20) | 0.92 | 0.85 (0.70-1.03) | 0.10 | 504 |
| Anorexia Nervosa | 0.68 (0.60 - 0.77) | 2.62E-09* | 1.84 (1.57-2.15) | 1.40E-14* | 1135 |
| Bulimia nervosa | 0.83 (0.72 - 0.97] | 2.22E-02 | 1.18 (1.00-1.39) | 0.05 | 771 |
| Autism Spectrum disorder | 0.97 (0.77 - 1.23) | 0.83 | 0.88 (0.70-1.1) | 0.26 | 356 |
| Panic disorder | 0.90 (0.81 - 0.99) | 3.60E-02 | 0.75 (0.68-0.83) | 5.3E-09* | 2092 |
| Obsessive compulsive disorder | 0.89 (0.89 - 1.05) | 8.96E-02 | 0.95 (0.94-1.08) | 0.46 | 1277 |
| PTSD | 0.96 (0.89 - 1.05) | 0.42 | 0.81 (0.74-0.88) | 4.96E-07* | 3130 |
| Personality disorder | 1.19 (1.05 - 1.36) | 7.90E-03 | 0.86 (0.76-0.97) | 0.02 | 1284 |
| Substance use disorder | 1.26 (1.06 - 1.49) | 7.69E-03 | 0.78 (0.66-0.9) | 8.0E-4* | 829 |
| Social anxiety disorder | 0.99 (1.06 - 1.49) | 0.81 | 0.85 (0.77-0.93) | 3.E4-4* | 2483 |
| Seasonal affective disorder | 0.97 (0.81 - 1.16) | 0.72 | 0.86 (0.72-1.02) | 0.08 | 629 |
| Neuroticism<sup>ø</sup> | 1.02 (1.01 - 1.04) | 2.15E-04* | 0.99 (0.98-1.00) | 0.16 | - |
| Extraversion<sup>ø</sup> | 0.99 (0.98 -1.01) | 0.34 | 1. (0.99-1.01) | 0.48 | - |
| All regressions adjused age and sex. *p<0.0028 significant after multiple testing correction; <sup>ø</sup>Effect per unit of raw score |  |  |  |  |  |

### Genetic factors

This section focuses on consenting to record linkage amongst those who provided a DNA sample. Necessarily, genetic analyses comparing participants who did or did not provide a biological specimen are not possible. Amongst those who provided a biological sample, 79% consented to PBS, implying a binomial variance of 0.79 (1- 0.79) = 0.17 on the observed scale. The GREML analysis suggested the presence of a genetic contribution to the likelihood of consenting to record linkage. The *SNP-based* heritability on the observed scale was 0.12 (S.E.= 0.03, p=1.1e-7, phenotypic variance on the observed scale∼0.16). We hypothesised that the genetic risk (operationalised as PRS) for the psychiatric traits identified above (e.g., bipolar disorder, anorexia or neuroticism) would be associated with differential consent for record linkage.

To assess specific genetic factors, we used logistic regressions to test for association between PRS for neuroticism, MDD, SCZ or EA. PRS were validated by first predicting the specific trait of interest. All PRS were predictive of their respective traits (**Table 3**). For example, neuroticism PRS was strongly associated with neuroticism score. Anorexia PRS (OR= 0.93 95% C.I.=[0.89 - 0.97]) and SCZ PRS (OR= 0.92 95% C.I.=[0.88 - 0.97]) were associated with lower odds of consenting to linkage. Conversely, MDD PRS was associated with higher odds of consent to record linkage (OR= 1.06 95% C.I.=[1.02 - 1.10]; **Table 3**). Using a GREML analysis, we estimated that MDD, SCZ and anorexia PRS accounted for around 12% of the SNP-based heritability of consent to record linkage (see Methods) which would account for around 1.4% of the total phenotypic variance on the observed scale. Notably all PRS were still significantly associated with agreeing to record linkage when jointly estimating their effects using GREML (MDDPRS beta=0.012 S.E.=3.7e-3; SCZPRS beta=-0.013 S.E.=4.9e-3; AnorexiaPRS=-9.2e-3 S.E.=3.8e-3).

**Table 3.**
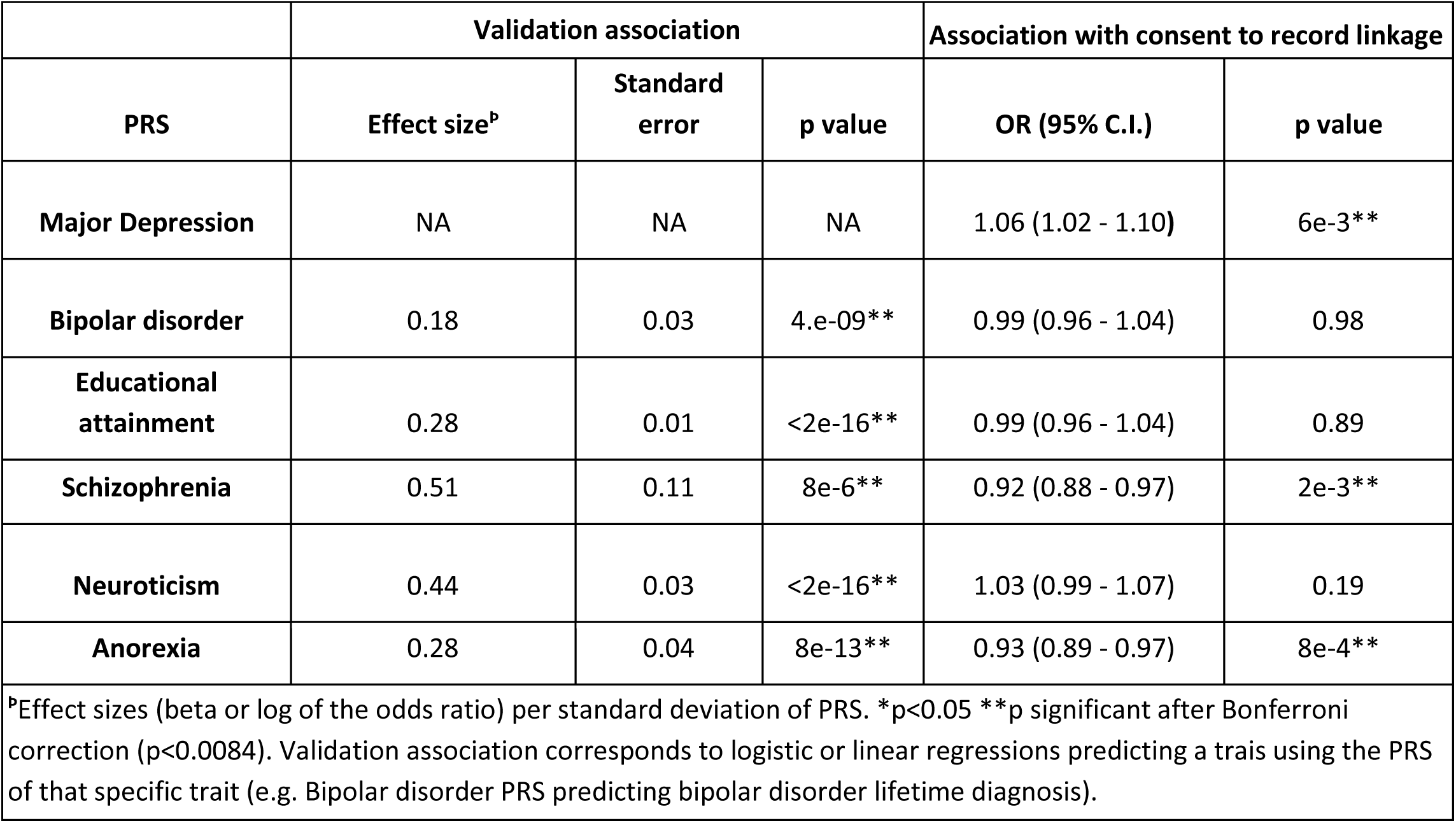
PRS validation and results of association with consent to record linkage.

|  |  |  |  |  |  |
| --- | --- | --- | --- | --- | --- |
|  | Validation association |  |  | Association with consent to record linkage |  |
| PRS | Effect size<sup>p</sup> | Standard error | p value | OR (95% C.I.) | p value |
| Major Depression | NA | NA | NA | 1.06 (1.02 - 1.10) | 6e-3** |
| Bipolar disorder | 0.18 | 0.03 | 4.e-09** | 0.99 (0.96 - 1.04) | 0.98 |
| Educational attainment | 0.28 | 0.01 | <2e-16** | 0.99 (0.96 - 1.04) | 0.89 |
| Schizophrenia | 0.51 | 0.11 | 8e-6** | 0.92 (0.88 - 0.97) | 2e-3** |
| Neuroticism | 0.44 | 0.03 | <2e-16** | 1.03 (0.99 - 1.07) | 0.19 |
| Anorexia | 0.28 | 0.04 | 8e-13** | 0.93 (0.89 - 0.97) | 8e-4** |
| <sup>p</sup>Effect sizes (beta or log of the odds ratio) per standard deviation of PRS. *p<0.05 **p significant after Bonferroni correction (p<0.0084). Validation association corresponds to logistic or linear regressions predicting a traits using the PRS of that specific trait (e.g. Bipolar disorder PRS predicting bipolar disorder lifetime diagnosis). |  |  |  |  |  |

## Discussion

This study investigated systematic differences between AGDS participants who consented to record linkage of prescription history and those who did not. We also investigated sociodemographic differences associated with the decision to donate a saliva sample for DNA analysis. Our study is motivated by the need to acknowledge and identify biases that could limit the generalisability of findings from prescription history data, and potentially lead to spurious associations.

Educational attainment has been reported to be a relevant factor for voluntary participation in medical research [9]. In fact, the AGDS has been shown to be a highly educated sample compared to the Australian population [1]. Here, we identified a moderate association between educational attainment and donating a biological sample, but the relationship between educational level and consent to record linkage did not reach statistical significance.

A recent study analysed genetic and demographic factors related to whether UK-Biobank participants shared their email for a follow up questionnaire. Women and individuals at high genetic risk for schizophrenia were less likely to share their email [8]. While we also identified that females were less likely to consent to record linkage, we found no association between consent to record linkage and a self-reporting of schizophrenia diagnosis. Nonetheless, we found a significant inverse association between SCZ PRS and consent to record linkage, which would align with the findings in the UK-Biobank and suggest our lack of phenotypic association to be due to lack of power from the low number of participants reporting schizophrenia or due to potential unreliability of a self-reported measure. Increased genetic risk for schizophrenia is likely related to higher basal suspiciousness, a heritable trait related to psychiatric disorders [20] which likely leads to a lower desire to record linkage.

Similar research has proposed that participants with higher neuroticism score and depression are less likely to participate in follow-up questionnaires. [9]. Participants with these characteristics may be more likely to experience feelings of anger, anxiety and irritability [21] when prompted with follow-up research surveys. In our study, individuals with higher neuroticism scores were more likely to consent to record linkage of their prescription history. We also report an association between neuroticism PRS and neuroticism score. However, the neuroticism PRS was not statistically associated with consent for record linkage. A similar result (i.e. phenotypic association, *valid* PRS but no association between PRS and consent to record linkage) for Bipolar disorder was observed. This observation could be explained by the fact that PRS does not capture all of the heritability for a trait and thus suffer from reduced power, but it may also imply that the relationship between neuroticism and consent to record linkage is mediated solely through the environmental component of neuroticism.

Participants reporting a lifetime diagnosis of bipolar disorder were more likely to consent to record linkage of their prescription history and less likely to provide a saliva sample for genotyping. Furthermore, genetic risk for depression was nominally associated with greater odds of consenting for record linkage, which is the reverse direction to the previous reports [9]. Participants who reported anorexia were less likely to consent to record linkage, but more likely to provide a biological specimen. This finding may be related to personality characteristics linked to anorexia, such as behaviors in relation to losing control [22]; consenting to access to the clinical prescription history could be violating the concept of having control [23]. However, this explanation fails to explain the positive association with donating a biological specimen.

Overall, the lack of consistency between demographic and clinical factors associated with consent for record linkage and donating a saliva sample, coupled with the results of other studies of participation and attrition, leads us to hypothesise that there is no single set of factors underlying attrition to different designs an settings of follow up studies. Evidence of differential factors in our study include the fact that previously reported negative predictors of participation, such as neuroticism and genetic risk for depression [9], were positively associated with consent to record linkage in our study; that educational attainment was associated with donating a saliva sample, but not with consent to record linkage and that a lifetime diagnosis of anorexia was associated with both record linkage and saliva donation but with opposing effects.

It is important to highlight some limitations of this study. The AGDS sample is ascertained for people who have a lifetime history of depression, the majority of whom have been prescribed antidepressants. These participants seem to be highly motivated to participate in the research and are highly enriched for help-seeking behaviours, making it a sensitised subset of the population. The diagnoses studied relied on participant self-report, which could lower the reliability of these diagnoses; however, as mental health disorders are highly impactful, a participants’ likelihood to accurately report the diagnoses communicated to them is high. Additionally, participants were informed of the time-period that would be covered by the record linkage and it is possible that participants who had not experienced a depressive episode in this time period may have been less likely to consent for record linkage (the previous 4.5 years from recruitment). It is also important to note that only Australian citizens and permanent residents have access to the PBS system and we did not collect information on legal citizenship or residentship status. Military personnel and their families do not use the MBS/PBS system. As such, non-residents and military personnel (and their families) would not be able to consent to PBS record linkage. Finally, within the genetic analyses, we excluded participants of non-European ancestry to avoid population stratification. For this reason, care should be taken when generalising these findings to populations with other ancestral backgrounds.

Overall our results suggest participation biases to be specific to the design and nature of the study. Our results show that not all observed differences are recapitulated through polygenic differences. These observations imply that using genetics is not a perfect tool to measure and adjust for unobserved biases. Conversely, subtle genetic differences may not be statistically identified using non-genetic data. For example, an association between SCZ PRS and consent to record linkage was not obvious from testing for association with a diagnosis for schizophrenia. The power to detect these differences using PRS should increase as discovery GWAS sample sizes increase. Thus, PRS will likely be a useful tool against attrition biases. Longitudinal studies with genotype data, such as the AGDS, enable us to identify these biases. Future studies leveraging the prescription data should consider these differences, and perform sensitivity analyses assessing whether their findings could be attributed to genetic risk for schizophrenia, depression or anorexia. Most current epidemiological studies do not adjust nor search for attrition biases at the genetic level. The possibility to do so is a strength of samples such as the AGDS. An example of such an approach is a recent study on depression identifying an unexpected positive genetic correlation between depression and cognitive traits was identified. Follow up analyses suggested these results to be explained by the fact that the AGDS sample is also highly educated whereas the education levels of the controls were more concordant with the Australian population [24]. We argue future analyses can be made more robust by identifying biases both on the phenotypic and genetic level, and either contextualising them as a potential limitation or performing sensitivity analyses adjusting for them.

## Data Availability

Summary data on effects described in this manuscript is available in the results section. GWAS summary statistics used in this study are publicly available and have been disclosed in the methods but are also available upon request. Code related to this study data analysis is available upon request from the authors. Access to the AGDS individual level data is restricted due to the ethical guidelines governing the study.

## Acknowledgements

Data collection for AGDS was possible thanks to funding from the Australian National Health & Medical Research Council (NHMRC) to NGM, NRW, SEM, IHB, EMB, PAL (GNT1086683) and Medical Research Future Fund (APP1200644). We thank our colleagues Richard Parker, Simone Cross, Scott Gordon and Lenore Sullivan for their valuable work coordinating all the administrative and operational aspects of the AGDS project. NRW thanks the support of NHMRC through grants 1113400 and 1173790. MER thanks the support of NHMRC and the Australian Research Council (ARC) through an NHMRC-ARC Dementia Research Development Fellowship (GNT1102821). SEM is supported in part by NHMRC investigator grant APP1172917. The views expressed are those of the authors and not necessarily those of the affiliated or funding institutions.

## Author Contributions

AIC and NGM designed the study. LG, SDT, LMGM and CY performed the analyses with supervision and guidance from AIC. EMB, PAL, NRW, SEM, IBH, and NGM implemented and supervised the data collection for the AGDS. LCC, LY, and MKL critically appraised the manuscript and gave input in the design and implementation of the analyses. LG, AIC and MER contributed to the first draft of the manuscript and all authors contributed to editing and drafting the manuscript prior to submission.

